# Determinants of male involvement in antenatal care at Palabek Refugee Settlement, Lamwo district, Northern Uganda

**DOI:** 10.1101/2023.02.13.23285867

**Authors:** Auma Irene, Nabaweesi Dinah, Orech Sam, John Bosco Alege, Allan Komakech

## Abstract

**Background:** In 2014, Uganda launched the National Male Involvement Strategy in Maternal and Child Health. In 2020, the District Health Management Information System report for Lamwo district, where Palabek Refugee Settlement is located, indicated a 10% male involvement in antenatal care (ANC). We investigated determinants of male involvement in ANC in Palabek Refugee Settlement to inform programs on improvement of male involvement in ANC in a refugee setting.

**Methodology:** We conducted a community-based cross-sectional analytical study among a proportionate sample of mothers in Palabek Refugee Settlement from October-December 2021. Using a standardized questionnaire, we collected information on demographics and the constructs of the socio-ecological model where consent was given. We summarized data in tables and figures. We used Pearson chi-square test to determine significance of independent variables at bivariate level. A multivariable logistic regression model was run for all variables found significant at bivariate analysis to determine association between the different independent variables and male involvement in ANC.

**Results:** We interviewed 423 mothers. The mean age of their male partners was 31 years, SD 7. Eighty-one percent (343/423) of male partners had formal education, with 13% (55/423) having a source of income and 61% (257/423) having access to ANC information during their pregnancy. The level of male involvement in ANC in Palabek Refugee Settlement was 39% (164/423). Male involvement in ANC was positively associated with access to information on ANC (AOR 3.0; 95%Cl: 1.7-5.4) and frequent couple discussion on ANC (AOR 10.1; 95%Cl: 5.6-18.0). However, it was negatively associated with distance ≥3km to the health facility (AOR 0.6 ;95%Cl: 0.4-1.0).

**Conclusions:** Approximately one in three male partners in Palabek Refugee Settlement were involved in ANC. Male partners who had access to information during ANC and those who had frequent discussions were more likely to get involved in ANC. Men who lived ≥3 kilometers from the health facility were less likely to be involved in ANC. We recommend intensified awareness creation on importance of male involvement in ANC and implementation of integrated community outreaches to reduce distance to the health facility.

## Introduction

Globally male involvement in maternal and child health remains low in both developed and developing countries (1).Some studies have indicated that few men get involved in antenatal care activities with less involvement being realized in the developing nations(2–4).

In low and middle-income countries male involvement in different aspects of maternal care is limited. An analysis of Demographic and Health Surveys across eight African countries indicated that less than half of all males were involved in antenatal care (ANC) with an average of 45.7% of men attending at least one ANC visit together with their partners (5).

A number of studies have suggested that male involvement in ANC is associated with benefits such as increased likelihood of adherence to HIV advice during pregnancy hence reduction of Mother to Child Transmission (MTCT) rates of HIV, improved utilization of maternal health services and reduced delays in the decision to seek health care(6–10).Despite its benefits, studies have also indicated that male involvement in ANC is affected by different individual, relational, institutional and societal related factors (11–15).

In Uganda, although the government launched the national male involvement strategy and guidelines in 2014, male involvement in ANC remains low. In a study conducted in Ibanda district in 2016, 50% of the women attending ANC did not have an attendant and the few who had them rarely had attendants as their husbands (16). In another study conducted in Uganda, male involvement in antenatal care was found to be only 6%(17).

The 2020 Lamwo district Heath Management Information system (HMIS) report indicated that only 10% of women from Palabek refugee settlement who attended antenatal care from July 2019 to June 2020 were accompanied by their partners for at least one antenatal care visit and less than 5% were accompanied by their spouses for four or more visits(18). However, male involvement in a refugee setting such as Palabek refugee settlement, which historically has family issues, is unknown.

The objectives of our study were to determine the factors associated with male involvement at Palabek refugee settlement to help inform programs aimed at improving male involvement in such a setting.

## Methods

### Study design and study setting

This was a community based analytical cross-sectional study which was conducted in Palabek refugee settlement in Lamwo District. In 2021 when the study was conducted, this settlement was made up of 9 zones that is Zone 1, Zone 2, Zone 3, Zone 4, Zone 5A, Zone 5B, Zone 6, Zone 7, and Zone 8.The settlement had a population of approximately 59,462 refugees, primarily from South Sudan(19). The settlement had three health facilities; Paluda Health Center III, Awich Health Center II and Akworo Health Center II. Other facilities within the host community where refugees seek health services are: Apyeta Health Center II and Palabek Ogili Health Center III. All these health facilities offer routine ANC services on week days with only emergency cases handled over the weekends.

### Study population

The study involved all females aged ≥18 years who were either six months and above expectant or gave birth in the previous one year before the study (October 2020 to October 2021).The team chose to interview female partners because we thought it would be difficult for the men to admit incase they were not involved in their partne’s ANC. In otherwords we thought the men would be baised and would not give the most accurate information. However, we thought the women would give us a more honest views on this.

### Sample size determination

The sample size was 384 calculated using the Kish Leslie formula based on the most conservative 50% estimated prevalence and a level of significance 5%. We further inflated this figure by 10% to account for non-response and recording errors during the study.

### Sampling procedure

We used probability proportionate sampling to calculate the numbers required for each zone. Data from Lamwo district HMIS report (2020) and data from village health teams’ registers was used to ascertain the number of pregnancies (≥6 months) and households with children ≤ 12 months. The second step involved conducting a convenience sampling of households with either an expectant mother or child below 1 year. To determine the direction of the zone where the interviews started, the research assistant stood at the center of the zone and threw a pen to the ground. The direction pointed by the lid became the starting point for interviews. Every expectant mother or mother who had a child below one year and gave consent was interviewed. When households in a particular direction got exhausted, the same process was repeated and the direction pointed by the pen lid was again used to determine the direction to be taken. This went on until the sample size was achieved.

### Data collection methods and tools

We conducted researcher-administered interviews using a standardized questionnaire with the help of six (6) research assistants. The first part captured information on different variables that affect male participation in ANC according to the constructs of the socio-ecological model. The second part assessed the level of men’s involvement in ANC.

### Study variables

The dependent variable was male involvement in antenatal care which was measured on a scale of 1-6 points with equal weights in the score adopted from Byamugisha et al index(20). The activities assessed using this scale included; whether the man makes joint plans with partner during pregnancy, whether the man attends ANC with partner, whether the man provides funding for ANC activities, whether the man helps in performing household chores, whether the man discusses with partner on issues occurring during ANC and whether the man discusses with partner’s healthcare provider on partner’s pregnancy. Each activity was given a score of one (1) if performed and zero (0) if not performed. A total score of 0-3 was considered non-involvement in ANC, while a score of 4-6 was considered as male involvement in ANC. The independent variables included the individual, relational, community and societal predictors of male participation in ANC.

### Data analysis

We analyzed data using Statistical Package for the Social Sciences (SPSS) version 22. At univariate level data was presented in frequencies and percentages. Age was expressed as a mean with its respective standard deviation. At bivariate level, associations between the dependent variable (male involvement in ANC) and the independent variables were determined using a Chi-Square test. The computed Chi-Square was compared to the critical value 0.05 level of significance at a 95% confidence interval. All the variables that were found to be significant (p<0.05) at the bivariate level were included in a binomial logistic regression model where crude odds ratios (ORs) and adjusted odds ratio and the 95% confidence intervals for each variable were calculated. The level of significance was equally set at P < 0.05.

### Ethics approval

Approval to carry out the study was obtained from the Clarke International University Research Ethical Committee No.**1.0, 2021-10-05**. Permission to collect data was sought from office of the Prime Minister (Palabek refugee desk). Informed written consent in the local language was sought from respondents. They were informed that their participation was voluntary, and their refusal would not result in any negative consequences. To protect the confidentiality of the respondents, each was assigned a unique identifier which was used instead of their names.

## Results

### Individual characteristics of male partners during a study to assess male involvement in ANC, Palabek Refugee Settlement, Lamwo district, Uganda, October-December, 2021

We interviewed a total of 423 females during this study. The mean age of the male partners was 31 years, SD 7. One hundred ninety-eight (47%) male partners were in the age group 28-37 years; majority (323, 76%) were married/cohabiting with their female partners. Three hundred forty-three (81%) had no formal education and most (257; 61%) had access to ANC information (Table 1)

**Table 1:** Individual characteristics of male partners.

| Variable | Frequency, n (%) |
| --- | --- |
| **Male partner's age** |  |
| 18-27 | 165 (39) |
| 28-37 | 198 (47) |
| 38-47 | 45 (11) |
| >48 | 15 (3) |
| **Marital status** |  |
| Married/cohabiting | 323 (76) |
| Unmarried | 100 (24) |
| **Level of education** |  |
| Formal education | 80 (19) |
| No formal education | 343 (81) |
| **Number of children with partner** |  |
| 0-2 | 194 (46) |
| 3-5 | 171 (40) |
| >6 | 58 (14) |
| **Source of income** |  |
| Yes | 55 (13) |
| No | 368 (87) |
| **Accessed ANC information during most recent pregnancy** |  |
| Yes | 257 (61) |
| No | 166 (39) |

### Level of male involvement in ANC, Palabek Refugee Settlement, Uganda, October-December, 2021

Of the 423 female respondents interviewed, 164 (39%) of their male partners were found to have been involved in ANC (Figure 1).

**Figure 1:**
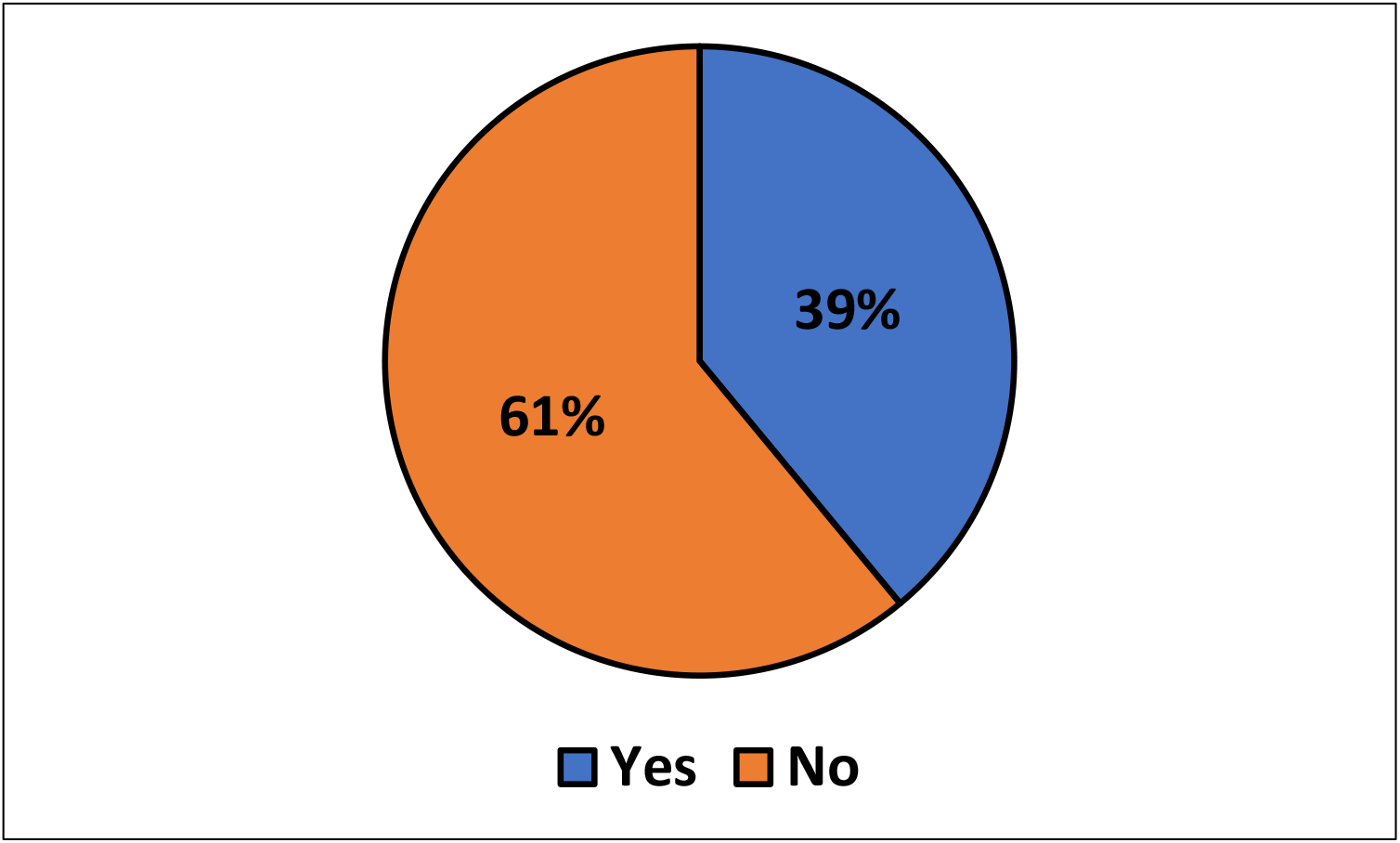
Male involvement in ANC, Palabek Refugee Settlement, Uganda, October-December, 2021

### Relational, institutional and societal characteristics of male partners during a study to assess determinants of male involvement in ANC, Palabek Refugee Settlement, Uganda, November 2021

Study findings indicated that 231 (55%) of respondents had frequent discussions of at least three times each week, 289 (68%) lived with their partners, 256(61%) lived with other family members while 234(55%) reported that peer influence affected male partner’s involvement in ANC (Table 2).

**Table 2:** Relational, institutional and societal characteristics of male partners.

| Variable | Frequency, n (%) |
| --- | --- |
| **Relational characteristics** |  |
| **Had frequent discussions on ANC during most recent pregnancy (n=423)** |  |
| Yes | 231 (55) |
| No | 192 (45) |
| **Lived with partner during most recent pregnancy (n=423)** |  |
| Yes | 289 (68) |
| No | 134 (32) |
| **Lived with other family members during most recent pregnancy(n=423)** |  |
| Yes | 256 (61) |
| No | 167 (39) |
| **Whether family members influenced partner's involvement in ANC (n=256)** |  |
| Yes | 154 (60 ) |
| No | 102 (40) |
| **Nature of influence of family members(n=154)** |  |
| Positive | 25 (16) |
| Negative | 129 (84) |
| **Whether peer influence affects male involvement in ANC (423)** |  |
| Yes | 234 (55) |
| No | 189 (45) |
| **Nature of peer influence (234)** |  |
| Positive | 51 (22) |
| Negative | 183 (78) |
| **Institutional characteristics** |  |
| **Whether health facilities promote male involvement (n=423)** |  |
| Yes | 383 (91) |
| No | 40 (9) |
| **Whether lack of male health workers affects male involvement (n=423)** |  |
| Yes | 7 (2) |
| No | 416 (98) |
| **Whether lack of privacy affects male involvement in ANC (n=423)** |  |
| Yes | 31 (7) |
| No | 392 (93) |
| **Whether long waiting hour affects male involvement in ANC (n=423)** |  |
| Yes | 315 (74) |
| No | 108 (26) |
| **Whether fear of HIV test affects male involvement in ANC (n=423)** |  |
| Yes | 217 (51) |
| No | 206 (49) |
| **Whether health worker's attitude affects male involvement in ANC (n=423)** |  |
| Yes | 56 (13) |
| No | 367 (87) |
| **Distance from partner's home affects male involvement in ANC (n=423)** |  |
| Less than 3km | 203 (48) |
| More than 3km | 220 (52) |
| **Societal factors** |  |
| **Whether cultural norms affect male involvement in ANC(n=423)** |  |
| Yes | 239 (57) |
| No | 184 (43) |
| **Whether there are structures that promote male involvement in ANC (n=423)** |  |
| Yes | 317 (75) |
| No | 106 (25) |
| **Whether there are government/health facility initiatives that affect initiatives male involvement in ANC (n=423)** |  |
| Yes | 231 (55) |
| No | 192 (45) |

Among the institutional characteristics, 315 (75%) of respondents thought that long waiting hours at the health facility during ANC clinics affects male involvement in ANC, 217 (51%) thought fear of HIV testing affects male involvement, 56 (13%) thought health worker’s attitude affects male involvement in ANC, while 220 (52%) of respondents reported that distance from their partner’s home to health facility was ≥3km

In regard to societal characteristics, 239 (57%) of respondents reported that cultural norms affect male involvement in ANC, 317 (75%) thought that there is an existence of structures that affect male involvement in ANC while 231 (55%) thought that government/health facility initiatives have an effect on male involvement in ANC (Table 2).

### Bivariate analysis showing relationship between male partner characteristics and involvement in ANC, Palabek Refugee Settlement, Uganda, November 2021

At bivariate analysis, we noted significant differences in male involvement concerning partner’s level of education (p= 0.001), source of income (p= 0.001), partner’s access to ANC information (p= 0.001), frequent discussion with female partner (p= 0.001), living with female partner during pregnancy (p= 0.001), average distance from partner’s home to the health facility (p= 0.004) and cultural norms (p= 0.035) (Table 3).

**Table 3:** Bivariate analysis of factors associated with male involvement in ANC in Palabek refugee settlement.

| Variable | Male involvement |  | X<sup>2</sup> | p-value |
| --- | --- | --- | --- | --- |
|  | Yes, n (%) | No, n (%) |  |  |
| **Individual predictors** |  |  |  |  |
| **Male partners age** |  |  | 5.6 | 0.134 |
| 18-27 | 59 (36) | 106 (64) |  |  |
| 28-37 | 77 (39) | 121 (61) |  |  |
| 38-47 | 24 (53) | 21 (47) |  |  |
| ≥48 | 4 (27) | 11 (73) |  |  |
| **Marital status** |  |  | 0.27 | 0.639 |
| Married | 123 (38) | 200 (62) |  |  |
| Un married | 41 (41) | 59 (59) |  |  |
| **Partner's level of education** |  |  | 11.0 | <0.001*** |
| No formal education | 18 (23) | 62 (77) |  |  |
| Formal education | 146 (43) | 197 (57) |  |  |
| **Number of children with partner** |  |  | 2.6 | 0.271 |
| 0-2 | 71 (37) | 123 (63) |  |  |
| 3-5 | 65 (38) | 106 (62) |  |  |
| 6 and above | 28 (48) | 30 (52) |  |  |
| **Source of income** |  |  | 16.5 | <0.001*** |
| Yes | 35 (64) | 20 (36) |  |  |
| No | 129 (35) | 239 (65) |  |  |
| **Whether partner accessed ANC information during most recent pregnancy** |  |  | 64.7 | <0.001*** |
| Yes | 139(54) | 118(46) |  |  |
| No | 25(15) | 141(85) |  |  |
| **Relational predictors** |  |  |  |  |
| **Whether there were frequent discussions on ANC** |  |  | 123.5 | <0.001*** |
| Yes | 145 (63) | 86 (33) |  |  |
| No | 19 (10) | 173 (90) |  |  |
| **Whether respondent lives/lived with partner during most recent pregnancy** |  |  | 16.5 | <0.001*** |
| Yes | 131 (45) | 158 (55) |  |  |
| No | 33 (25) | 101(75) |  |  |
| **Whether respondent lives/lived with other family members during most recent pregnancy** |  |  | 0.003 | 1.000 |
| Yes | 99 (39) | 157 (61) |  |  |
| No | 65 (39) | 102 (61) |  |  |
| **Whether peers influence affects male involvement in ANC** |  |  | 0.02 | 0.920 |
| Yes | 90 (38) | 144 (62) |  |  |
| No | 74 (39) | 115 (61) |  |  |
| **Institutional predictors** |  |  |  |  |
| **Whether health facilities promote male involvement in ANC** |  |  | 1.43 | 0.240 |
| Yes | 152 (40) | 231 (60) |  |  |
| No | 12 (30) | 28 (70) |  |  |
| **Whether absence of male staff affects male involvement in ANC** |  |  | 0.31 | 0.711 |
| Yes | 2 (29) | 5 (71) |  |  |
| No | 162 (39) | 254 (61) |  |  |
| **Whether lack of privacy affects male involvement in ANC** |  |  | 0.15 | 0.709 |
| Yes | 11 (35) | 20 (65) |  |  |
| No | 153 (39) | 239 (61) |  |  |
| **Whether long waiting time affects male involvement in ANC** |  |  | 0.51 | 0.494 |
| Yes | 119 (38) | 196 (62) |  |  |
| No | 45 (42) | 63 (58) |  |  |
| **Whether fear of HIV test affects male involvement in ANC** |  |  | 0.15 | 0.842 |
| Yes | 83 (38) | 134 (62) |  |  |
| No | 81 (39) | 125 (61) |  |  |
| **Whether attitude of health workers affects male involvement in ANC** |  |  | 1.93 | 0.187 |
| Yes | 17(30) | 39 (70) |  |  |
| No | 147(40) | 220 (60) |  |  |
| **Whether distance from partner's home to health facility affects male involvement in ANC** |  |  | 8.63 | **0.004\*** |
| Less than 3km | 64(32) | 139(68) |  |  |
| More than 3km | 100(45) | 120(55) |  |  |
| **Societal predictors** |  |  |  |  |
| **Whether cultural norms affect male involvement in ANC** |  |  | 4.6 | **0.035\*** |
| Yes | 82(34) | 157(66) |  |  |
| No | 82(45) | 102(55) |  |  |
| **Whether structures that affect male involvement in ANC** |  |  | 2.7 | 0.108 |
| Yes | 130 (41) | 187 (59) |  |  |
| No | 34 (32) | 72 (68) |  |  |
| **Whether there are policy factors/initiatives that affect male involvement in ANC** |  |  | 1.70 | 0.229 |
| Yes | 96 (41) | 135 (59) |  |  |
| No | 68 (35) | 124 (65) |  |  |

**Table 4: Multivarate analysis of factors associated with male involvement in ANC in Palabek Refugee Settlement, Uganda, November 2021.**

At multivariate analysis, we noted that access to ANC information by male partners (AOR 3.0; 95%CI: 1.7-5.4), having discussions on ANC (AOR 10.1; 95%CI: 5.6-18.0) and living more than 3kms from the health facility (AOR 0.037, 95%CI: 0.6-1.0) were significantly associated with male involvement (Table 4).

**Table 4:** Multivariate analysis of factors associated with male involvement in ANC in Palabek refugee Settlement.

| Variable | Crude odds ratio (95% CI) | p-value | Adjusted odds ratio (95% CI) | p-value |
| --- | --- | --- | --- | --- |
| **Level of education** |  |  |  |  |
| No formal education | 1.0 |  | 1.0 |  |
| Formal education | 0.6(0.3-1.3) | 0.188 | 0.6(0.3-1.3) | 0.178 |
| **Economic status** |  |  |  |  |
| Did not have a source of income | 1.0 |  | 1.0 |  |
| Had a source of income | 1.8(0.9-3.5) | 0.102 | 1.8(0.9-3.6) | 0.082 |
| **Access to information** |  |  |  |  |
| No | 1.0 |  | 1.0 |  |
| Yes | 3.0(1.7-5.4) | <0.001 | 3.0 (1.7-5.4) | **0.001\*\*\*** |
| **Discussion on ANC** |  |  |  |  |
| No | 1.0 |  | 1.0 |  |
| Yes | 9.8(5.5-17.4) | <0.001 | 10.1 (5.6-18.0) | **<0.001\*\*\*** |
| **Living with partner** |  |  |  |  |
| No | 1.0 |  | 1.0 |  |
| Yes | 1.6 (0.9-2.9) | 0.102 | 1.6 (0.9-2.9) | 0.107 |
| **Average distance to health facility** |  |  |  |  |
| Less than 3Km | 1.0 |  | 1.0 |  |
| 3km and above | 0.6 (0.4-1.0) | 0.045 | 0.6 (0.4-1.0) | **0.037\*** |
| **Whether cultural norms affect male involvement in ANC** |  |  |  |  |
| No | 1.0 | 1.0 |  |  |
| Yes | 0.8 (0.5-1.4) | 0.511 | 0.8 (0.5-1.4) | 0.420 |

Key: \*=p-value<0.05, \*\*=p-value<0.01, \*\*\*=p-value<0.001

## Discussion

The study findings indicated that one in three men were involved in ANC. Access to ANC information and frequent couple discussions on ANC were positively associated with male involvement. On the other hand, longer distance of ≥3km to the health facility had a negative association with male involvement in ANC.

The study findings indicated that one in three men were involved in ANC. This is higher than the level of male involvement stipulated by some studies conducted in different parts of Africa including Uganda & Zambia(17,21). This difference is probably because in the comparison studies level of male involvement was measured by a man’s attendance of ANC together with partner while in the current study level of male involvement was measured by a man’s participation in six different activities. In this study a man’ s participation in at least four of the activities meant that the man was involved while participation in ≤3 or less activities meant that the man was not involved in ANC. The study finding also indicated that level of male involvement in the current study was lower than the level of involvement indicated in other studies conducted in Tanzania, Ghana, Kenya and India (22–25). The WHO report on refugee and migrant health for 2022 (26) supports the above findings. This report indicated that utilization of health services among refugees is restricted due to social economic, political barriers, cultural difference and institutional discrimination. It is therefore inevitable that optimal utilization of ANC services which would be improved by male involvement is also affected by such barriers.

The study findings indicated that men who had access to ANC information were three times more likely to be involved in ANC than their counter parts. In proposition of above finding, the resource package on engaging men during pregnancy and child birth UNFPA, UN women and European Union (27) suggests that health education for men is very important in improving male involvement in ANC. These study findings are also similar to findings of different studies conducted in Africa including Dodoma region of central Tanzania and Ambo town, Ethiopia (22,28) where male involvement was higher among men who have access to information. The men who have access to information are more likely to be involved in ANC probably because access to information leads to improved level knowledge on the importance of their involvement. It is important that a number of awareness creation interventions such as use of Information Education and communication (IEC) materials, modeling through peers, mass media and use of theatre as a tool for health promotion should be implemented.

This study finding indicated that slightly above half of male partners engaged in frequent communication on ANC during their partners’ most recent pregnancy. The study findings also indicated that female partners who reported that they had frequent discussions on ANC with their spouses were up to 10.1 times more likely to have their partners involved in ANC than their counter parts who reported that they did not have such discussions. This is probably because when couples communicate on matters concerning ANC, they are likely to learn the needs of the expectant partners and also agree on how the male partner can support during this period. These findings are in agreement with the findings conducted in Kyela district, Mbeya Tanzania and Dodoma region of Tanzania where partners who had frequent discussion were likely to realized higher odds of male involvement in ANC(3,22).

The study findings indicated that approximately half of male partners lived at a distance of less than 3km from the nearest health facility. The study findings further indicated that the men who lived more than 3km from the nearest health facility were 0.4 times less likely to get involved in ANC than their counter parts who lived less than 3km to the nearest health facility. UNICEF in its community based program for bringing critical services to the world’s hardest-to-reach children and mothers suggested that one of the greatest barriers for service utilization is the distance to the health facilities (29). This barrier definitely affects male involvement in ANC like it affects access to other health services. These findings are similarly to the study findings for a study conducted in Sekondi region of Ghana(14).The men who live closer to the health facilities are more likely to be involved in ANC probably because they are less likely to incur transport cost as a couple to reach the health facilities. Such men are also likely to spend a shorter time getting to the health facilities and can receive services earlier then go back for their other businesses unlike their counter parts. The study findings are however contrary to findings of a Study conducted in South Ethiopia (30) which suggested that that women who walked less than 5 km distance to the nearest health facility had lower odds of having their partners involved in ANC than their counterparts who walked more than 5 km. It is important to accelerate strategies such as conducting regular integrated outreaches to ensure that services are taken closer to the community members which would in turn lead to increased male involvement in ANC.

### Strengths and limitations of the study

Our study had some strengths. We undertook a holistic approach using six variables to assess the level of male involvement. This makes it different from other studies that majorly employed one variable (accompanying partner for ANC clinics) to measure the level of male involvement. Secondly, we used constructs of the Social Ecological Model to develop our questionnaire. The use of such a standardized tool ensures validity, reliability and objectivity of our study findings. This was the first study on male involvement in ANC to be conducted in a refugee setting in Uganda. It therefore provides an opportunity for generalization of findings to similar refugee communities.

Our study also had some limitations. First our study was conducted in the refugee settlement where movement is restricted so it may be difficult to generalize findings for the stable communities since these contexts are completely different. Furthermore, we ascertained determinants of male involvement in the most recent pregnancy. The level of involvement for earlier pregnancies could have been different with different influencing factors.

## Conclusion and recommendations

The study findings indicated that one in every three men at Palabek refugee settlement were involved in ANC. Two factors that is access to information on ANC and frequent couple communication were found to be positively associated with level of male involvement. On the other hand, distance (> 3km) to the health facility had was negatively association with male involvement in ANC.

We therefore recommend that the government and UNHCR should accelerate community sensitization on the importance of male involvement and the role of male partners during pregnancy.

The government of Uganda and UNCHR should emphasize implementation of the modeling approach where men with high involvement index are modeled to become change agents.

Acceleration of strategies such as regular integrated outreaches would ensure that services are taken closer to the community members. This would in turn lead to increased male involvement in ANC. Finally, we recommend that a qualitative study should be conducted to assess in-depth community views and insights on determinants of male involvement in ANC and ways of bridging identified gaps.

## Data Availability

All data produced in the present study are available upon reasonable request to the authors

## List of abbreviations

ANC: Antenatal care
AOR: Adjusted odds ratio
EU: European Union
IEC: Information Education and communication
Km: Kilometer
MCTC: Mother -to -child transmission of HIV
OPM: Office of the Prime Minister
SPSS: Statistical Package for Social Sciences
UNFPA: United Nations Population Fund
UNHCR: United Nations High Commission for Refugees
UNICEF: United Nations Children’s Fund
UN women: United Nations Women
WHO: World Health Organization

## Declarations

### Ethics approval and consent to participate

All the protocols for this project were reviewed and approved by Clarke International University (CIU) Research Ethical Committee No.1.0, 2021-10-05 in accordance with the Declaration of Helsinki and CIU research guild lines. Permission to collect data from the settlement was sought from office of the Prime Minister (Palabek refugee desk). We also sought verbal informed consent in the local language from the participants as we had used phone interviews and the verbal informed consent procedure was the most suitable. They were informed that their participation was voluntary, and their refusal would not result in any negative consequences. To protect the confidentiality of the respondents, each was assigned a unique identifier that was used instead of their names. All methods were carried out in accordance with relevant guidelines and regulations.

### Consent for publication

Not applicable

### Availability of data and materials

The datasets used and/or analyzed during the current study are available from the corresponding author on reasonable request. The data set and questionnaires used and/or analyzed during the current study are available on Mendeley repository.

### Authors’ contributions

IA did the conceptualization of the study idea, data analysis, writing, and editing of the manuscript. DN, SO, JBA and AK were involved in the conceptualization of the study idea. AK and JBA guided the writing and reviewing of the manuscript and were involved in the conceptualization of the study idea, writing, editing, and reviewing of the manuscript. All authors read and approved the final manuscript.

## Acknowledgements

We thank the respondents for their participation in the study. We also thank the Office of the Prime Minister (OPM) Palabek Refugee Desk for the administrative clearance provided before conducting this study. Finally, we thank the research assistants for doing data collection.

